# LITE SABR M1: a Phase I Trial of Lattice Stereotactic Body Radiotherapy for Large Tumors

**DOI:** 10.1101/2021.06.05.21257825

**Authors:** Sai Duriseti, James Kavanaugh, Jeff Szymanski, Yi Huang, Franco Basarabescu, Aadel Chaudhuri, Lauren Henke, Pamela Samson, Alexander Lin, Clifford Robinson, Matthew B Spraker

## Abstract

**Purpose:** Stereotactic body radiotherapy (SBRT) is an attractive treatment option for patients with metastatic and/or unresectable tumors, however its use is limited to smaller tumors. Lattice is a form of spatially fractionated radiotherapy that may offer safe delivery of SBRT for bulky tumors. We previously described Lattice SBRT, which delivers 20 Gy in 5 fractions with a simultaneous Lattice boost to 66.7 Gy. The goal of this study was to prospectively evaluate the acute toxicity and quality of life (QoL) of patients with large tumors (> 5 cm) treated with Lattice SBRT.

**Methods:** This was a single-arm phase I trial conducted between October 2019 and August 2020. Patients with tumors >4.5 cm were eligible. Lattice SBRT was delivered every other day.. The primary outcome was the rate of 90-day treatment-associated (probably or definitely attributable) grade 3+ acute toxicity by Common Terminology Criteria for Adverse Events (CTCAE) version 5.0 criteria. Other outcomes included changes in patient reported toxicity and QoL inventories, GTV, and peripheral blood cytokines.

**Results:** Twenty patients (22 tumors) were enrolled. Median GTV was 783 cc (range: 64-3713 cc). Fifty percent of tumors were in the thorax, 45% abdomen/pelvis, and 5% extremity. There was no treatment-associated grade 3+ toxicity.. There was one case of grade 4 toxicity possibly associated with Lattice SBRT.

**Conclusions:** This phase I study met its primary endpoint of physician reported safety. An ongoing phase II clinical trial of Lattice SBRT will evaluate late safety and efficacy of this novel technique.

## Introduction

Hypofractionated radiotherapy is an appealing treatment for patients with metastatic or unresectable tumors in need of palliative radiotherapy.^1^ Stereotactic body radiotherapy (SBRT) is a form of hypofractionated radiotherapy increasingly used in this setting as it may improve local control and better palliate symptoms compared to conventional fractionation.^2,3^ SBRT does face limitations when considered for large tumors. Locoregional control and toxicity are both thought to be worse for larger tumors treated with SBRT compared with smaller tumors.^4,5^ Toxicity has also been reported to be worse for larger tumors treated with SBRT.^6–8^ Consequently, consensus guidelines support use of SBRT for tumors 5 cm or smaller.^9^

Spatially-fractionated radiotherapy purports to allow for safe, hypofractionated, dose-escalated radiotherapy for large tumors.^10^ Spatially-fractionated radiotherapy can be delivered in a grid-like pattern (i.e. GRID) using 3D conformal planning techniques. It can also be delivered using an intensity modulated technique (i.e. Lattice radiotherapy) that simultaneously delivers a conformal lower dose of radiation to a planning target volume (PTV) with a simultaneous integrated boost to a pre-defined geometry of spherical sub-volumes confined to the gross tumor volume (GTV).^11^ Both GRID and Lattice radiotherapy have been used as definitive treatment, as a boost after conventional radiation, and de novo or for reirradiation. While the literature has been limited to retrospective studies, GRID and Lattice have associated with significant clinical responses and limited toxicities in large and/or radioresistant tumors, including non-small cell lung cancer and sarcomas.^12–15^

We previously described a spatially fractionated SBRT technique, Lattice SBRT, that delivers 20 Gy in 5 fractions with a 66.7 Gy simultaneous integrated Lattice boost using volumetric modulated arc therapy (VMAT).^16^ Here, we present the results of LITE SABR M1: a single-arm phase I study of Lattice SBRT for large tumors (NCT04133415).

## Materials/Methods

### Study Design and Patients

This study was a prospective, single-arm, single-institution phase 1 trial. It was approved by the local institutional review board and all patients provided written informed consent. Eligible patients were at least 18 years of age, had a histologically confirmed cancer diagnosis, had a tumor of at least 4.5 cm in largest axial diameter, and Eastern Cooperative Group (ECOG) performance status of 2 or less. All patients received a multidisciplinary recommendation for radiotherapy for palliation of symptoms associated with the tumor target, asymptomatic progression, or as definitive treatment for locally-advanced unresectable and/or metastatic tumors. Patients who received or planned to receive systemic therapy before or after Lattice SBRT were eligible, although a 2 week washout was recommended before or after Lattice SBRT. Concurrent systemic therapy was not allowed. Patients were ineligible if they previously received radiotherapy to the same area, if the tumor needed emergent surgical intervention, if they were pregnant, or had uncontrolled HIV with CD4 below 350 ^cells^/_µl_ or on certain anti-retroviral therapies.

### Treatment Planning and Delivery

Our treatment planning and quality assurance (QA) process has been detailed previously, and a treatment planning guide is available.^16^ Briefly, computed tomography (CT) simulation was completed with 3 mm slices and four-dimensional (4D) CT was used to assess tumor motion for all patients with thoracic and abdominal tumors above the iliac crest. Gated radiotherapy was used if tumor motion was more than 5 mm in a single direction. The planning process is fully described in the companion paper by Kavanaugh et al., but it is briefly summarized as follows. The treating radiation oncologist created a gross tumor volume (GTV) and fusion of prior diagnostic imaging was allowed. They then created a 5–10 mm isotropic expansion of the GTV to form the PTV_2000. Using the GTV, a dosimetrist and/or physicist created a PTV_6670 (i.e. the Lattice boost volume) and other Lattice planning volumes (i.e. PTV_Avoid). Volumetric-modulated arc therapy (VMAT) Lattice SBRT plans were generated in the Eclipse Treatment Planning System (Varian, Palo Alto, CA). The protocol required that all plans meet consensus 5-fraction SBRT organ at risk (OAR) constraints published by the American Association of Medical Physicists.^17^ If a plan could not meet the OAR constraints, the PTV_6670 was modified. Standard clinical SBRT QA protocols used for all plans are described in the companion paper.

Lattice SBRT was delivered every other day for a total of 5 fractions. Patients could receive Lattice SBRT to multiple sites on alternating days if the radiotherapy plans did not overlap. Patients could also receive other conventional palliative radiotherapy courses to other lesions during the study period as directed by the treating physician. On-board imaging was required to verify positioning for all patients. Cone-beam CT (CBCT) and orthogonal kV films or fluoroscopic imaging were obtained immediately prior to treatment for tumor position verification. Structures were created and displayed on the CBCT from critical dose lines to assist with OAR position verification at the time of treatment.

### Follow Up, Physician, and Patient Reported Outcomes

Patients were assessed for acute toxicity through 90 days after completion of therapy and were evaluated in clinic at baseline, once during therapy, and at 14, 30, and 90 days. Physician-reported and patient-reported outcomes (PROs) were evaluated at each visit using CTCAE version 5.0 and PRO-CTCAE, respectively. PROMIS anxiety (Item Bank v1.0 – Emotional Distress – Anxiety), physical global health (v1.2, physical 2a), depression (v1.0, short form 4a), pain interference (v1.0, short form 4a), and physical function (v2.0, short form 10a). were assessed at each visit.^18,19^

### Statistical Analysis

The primary endpoint of the study was the rate of treatment-associated (probably or definitely attributable) grade 3 or greater CTCAE v5.0 toxicity within 90 days of completion of Lattice SBRT. Early stopping of the trial was based on a Pocock-type stopping boundary. We assumed an expected treatment emergent severe adverse event rate of 30% with an unacceptable rate of higher than 40%. The trial opened on 10/31/2019 with an enrollment goal of 10 patients. The trial was expanded since accrual was robust and to allow for better detection of possible severe acute toxicities prior to opening a planned phase II trial. The study was expanded to 15 patients on 4/23/2020 and then to 20 patients on 6/17/2020. Trial accrual by month is exhibited in Supplementary Figure 1. The trial completed enrollment on 08/11/2020.

Secondary objectives included assessment of patient-reported toxicity and quality of life (QOL) at baseline and in follow up. Toxicity was assessed using PRO-CTCAE according to site-specific inventories. A composite score was calculated for each pertinent symptom for thoracic tumors and abdominal/pelvic tumors all time points. The composite score combines the severity, frequency, and interference elements of each toxicity into a single score for efficient reporting.^20^ PROMIS anxiety (Item Bank v1.0 – Emotional Distress – Anxiety), global health – physical (2a, v1.2), pain interference (Short Form 4a), and physical function (Short Form 10a) was used to asses PROMIS QOL scores.^21^ PROMIS inventories were scored using the Health Measures Scoring Service (https://www.assessmentcenter.net/ac_scoringservice) and the median T score was calculated for each patient for each available time point.

An exploratory analysis of tumor imaging response after Lattice SBRT was completed. All available follow up images at least 30 days from completion of treatment were imported into the Eclipse TPS (Varian, Palo Alto) and fused with each patient’s CT simulation series. Using the originally contoured GTV_2000 as a benchmark, the treated lesions were contoured on each available diagnostic CT scan and assessed for percent change from baseline by volume.

An additional exploratory analysis of cytokine/chemokine changes was performed on patient plasma collected at baseline, post-treatment, and at 14- and 30-day follow up. We hypothesized that Lattice SBRT would be associated with significant changes in cytokine/chemokines related to the immune response to radiotherapy. The methods for specimen collection, testing, and analysis are available in the supplementary materials.

## Results

### Patient and Disease Characteristics and Disease-specific Outcomes

Between October 2019 and August 2020, 20 patients underwent Lattice SBRT. Two patients were treated to two sites each, for a total of 22 tumors treated. The CONSORT diagram for the study is shown in Figure 1. Patient and tumor characteristics are shown in Table 1. The population was 50% female, median age was 67 years (range: 31 to 86), and median ECOG status was 1 (range 0 to 2). Thirteen patients (65%) identified as White, 6 patients (30%) identified as Black, and 1 patient (5%) identified as Asian. Nine patients had soft tissue sarcomas (45%), 7 non-small cell lung cancers (35%), 1 thymic carcinoma (5%), 1 malignant mesothelioma (5%), 1 endometrial adenocarcinoma (5%), and 1 colonic adenocarcinoma (5%). Median GTV was 579.2 cc (range: 54.2–3713.5 cc) and median largest diameter of the tumor was 11.1 cm (range: 5.6–21.4 cm). Fifteen patients (75%) had undergone at least 1 course of chemotherapy and 3 patients (15%) received radiotherapy to other sites prior to receiving Lattice SBRT. Eight patients (40%) received chemotherapy after completion of Lattice SBRT. The median time from CT simulation to first fraction was 12 days (range: 6–21 days). The median length of the course of treatment was 11 days (range 9–23 days). Three patients (15%) underwent Lattice SBRT as their only form of therapy. At 90-day follow up, 80% of treated patients were alive for evaluation.

**Table 1:** Patient clinical information, disease information, gross tumor volume (GTV), time from last day of systemic therapy prior to Lattice SBRT (days), time from completion of Lattice SBRT to initiation of subsequent systemic therapy (days)

| PARTICIPANT | ECOG | INDICATION | SEX | SITE | HISTOLOGY | GTV (CC) | TIME PRIOR TO LATTICE | TIME AFTER LATTICE |
| --- | --- | --- | --- | --- | --- | --- | --- | --- |
| 1* | 2 | Palliation | F | Thorax | NSCLC, poorly-differentiated | 1784.2 | -- | -- |
| 2* | 1 | Palliation | M | Thorax | Solitary fibrous tumor | 616.9 | 281 | -- |
| 3* | 1 | Palliation | F | Thorax | NSCLC, adenocarcinoma | 551.4 | -- | 6 |
| 4* | 1 | Oligoprogression | M | Thorax | Leiomyosarcoma | 64.3 | 49 | -- |
| 5* | 0 | Oligoprogression | M | Thorax | NSCLC, squamous cell carcinoma | 318.4 | 126 | 143 |
| 6 | 2 | Definitive | M | Abdomen | Undifferentiated pleomorphic sarcoma | 644.8 | 52 | 39 |
| 7* | 2 | Palliation | M | Thorax | Neuroendocrine tumor | 606.9 | 396 | -- |
| 8 | 1 | Palliation | F | Thorax | NSCLC, squamous cell carcinoma | 251.1 | -- | -- |
| 9* | 1 | Palliation | M | Abdomen | Round cell sarcoma | 67.0 | 14 | -- |
| 10* | 0 | Oligoprogression | M | Thorax | Rhabdomyosarcoma, pleomorphic type | 54.2 | 58 | 1 |
| 11*+ | 1 | Definitive | F | Abdomen | Leiomyosarcoma | 3713.5 | -- | -- |
|  |  |  |  | Extremity |  | 1426.3 |  |  |
| 12*⊠ | 2 | Palliation | F | Thorax | NSCLC, adenocarcinoma | 206.9 | -- | 24 |
|  |  |  |  | Abdomen |  | 316.7 |  |  |
| 13* | 2 | Palliation | F | Pelvis | Rhabdomyosarcoma, embryonal type | 1666.6 | 21 | -- |
| 14* | 1 | Oligoprogression | F | Pelvis | NSCLC, poorly-differentiated | 208.3 | 37 | -- |
| 15 | 1 | Definitive | F | Thorax | Thymoma | 815.1 | 77 | -- |
| 16* | 1 | Palliation | F | Thorax | Mesothelioma | 1486.4 | 48 | 4 |
| 17* | 1 | Palliation | M | Abdomen | Endometrial, adenocarcinoma | 131.7 | 342 | 11 |
| 18* | 2 | Palliation | F | Pelvis | Colon adenocarcinoma | 3616.2 | 29 | -- |
| 19* | 2 | Palliation | F | Pelvis | Desmoplastic small round cell tumor | 462.7 | 15 | 16 |
| 20* | 2 | Oligoprogression | M | Abdomen | Leiomyosarcoma | 1573.2 | 47 | -- |

NSCLC = Non-small cell lung cancer
\* Patients with metastatic disease
+ Patient 11 had 2 sites of disease, her primary retroperitoneal tumor and a single site of metastatic disease in the extremity.
⊠ Patient 12 had 2 sites of disease, her primary tumor in her right lung and a single site of metastatic disease in the right adrenal gland.

**Figure 1:**
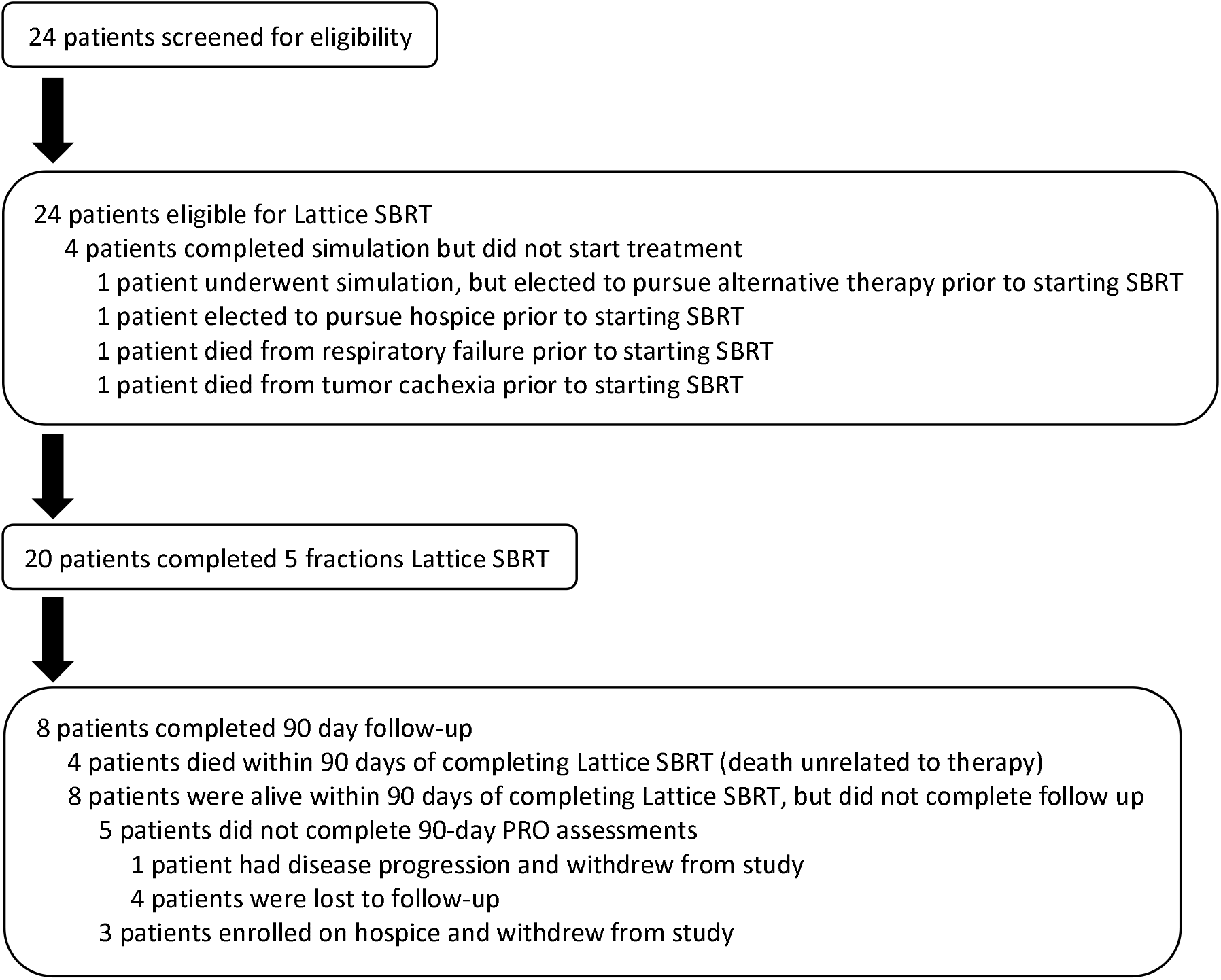
LITE SABR M1 Consort diagram

### Toxicity

All patients successfully completed the prescribed Lattice SBRT course. Cumulative acute toxicities within 90 days of therapy are shown in Table 2. Only one patient (Patient 11) had grade 3 or greater toxicity possibly related to Lattice SBRT. She underwent Lattice SBRT to a 3713.5 cc primary retroperitoneal leiomyosarcoma and a 1426.3 cc right thigh metastasis. She had known ureteral involvement from her retroperitoneal tumor and had percutaneous nephrostomy tubes placed before starting treatment. After 2 fractions of Lattice SBRT to her retroperitoneal tumor and 1 fraction to her extremity metastasis, she was admitted for grade 4 urosepsis requiring pressor support, antibiotics, and nephrostomy tube exchange. She resumed treatment to her retroperitoneal and extremity tumors 12 and 14 days after the prior fraction to each site, respectively. She also had grade 3 transaminitis concurrent with her sepsis and grade 2 diarrhea that resolved after treatment. On plan review, a segment of the intra-renal portion of the tube was inside of the 2000 cGy isodose line, and the soft tissue traversed by the tube was within the 1000 cGy isodose line.

**Table 2:** <u>Attribution of toxicity for patients treated with Lattice SBRT.</u> The total number of toxicities graded per CTCAE version 5.0 criteria are shown.

|  | Unrelated |  |  | Unlikely |  |  | Possibly |  |  |
| --- | --- | --- | --- | --- | --- | --- | --- | --- | --- |
| Grade | 2 | 3 | 4 | 2 | 3 | 4 | 2 | 3 | 4 |
| Total Events | 53 | 14 | 1 | 6 | 2 | 1 | 1<sup>*+</sup> | 2<sup>+</sup> | 1<sup>+</sup> |

\* Patient 2 had increased dyspnea and work of breathing about 60 days after completion of therapy, clinically consistent with radiation pneumonitis. He was treated with prednisone and had resolution of his symptoms.
+ Patient 11 required hospitalization for urosepsis after 2 fractions of Lattice SBRT to her abdominal tumor. She had Grade 3 transaminitis, attributed to her sepsis physiology. She also had Grade 2 diarrhea that self-resolved, however given Lattice SBRT delivery to the abdomen, an acute radiotherapy toxicity could not be excluded.

Patient 2 experienced grade 2 pneumonitis possibly associated with Lattice SBRT. The patient was treated with Lattice SBRT to a 616.9 cc right, anterior mediastinal metastasis from solitary fibrous tumor. Sixty days after completion of therapy he reported increased dyspnea on exertion consistent with radiation pneumonitis. No imaging was obtained. He was treated with prednisone and his symptoms resolved. There were no deaths associated with Lattice SBRT.

### Patient reported outcomes

Prior to treatment, 95% of the total cohort (19 of 20 eligible) completed baseline PRO evaluation. At 14 days post-treatment, 84% of eligible patients (16 of 19 eligible) completed post-treatment evaluation, with 3 patients declining PRO follow up at this time point, and 1 patient unavailable due to enrollment on hospice. At 30 days post-treatment, 56% (11/18) completed post-treatment evaluation, with 7 patients declining PRO follow up at this time point, 1 unavailable due to enrollment on hospice, and 1 deceased. At 90 days post-treatment, 62% (8/13) completed post-treatment evaluation, with 5 declining PRO follow up at this time point, 3 unavailable due to enrollment on hospice, and 4 deceased within 90 days follow up.

The proportions of patients thoracic and abdominal/pelvic tumors with grade 0-1 and grade 2+ composite PRO-CTCAE scores at baseline and 14 days after therapy are shown in Figure 2. In patients with thoracic targets, the proportion of patients with grade 2+ shortness of breath decreased from baseline (33%) to 14 days (11%) after Lattice SBRT. The proportion of patients with grade 2+ slightly increased for vomiting (11% to 22%) and was unchanged for pain and fatigue from baseline to 14 days. In patients with abdominal or pelvic tumors, there was a decrease in the proportion of patients with grade 2+ pain (86% to 57%), GI/GU symptoms (71%-43%), and anxiety (43% to 14%). The proportion of patients withfatigue remained constant (86%) from baseline to 14 days. All PRO-CTCAE data is shown in Supplementary Table 1.

**Figure 2.**
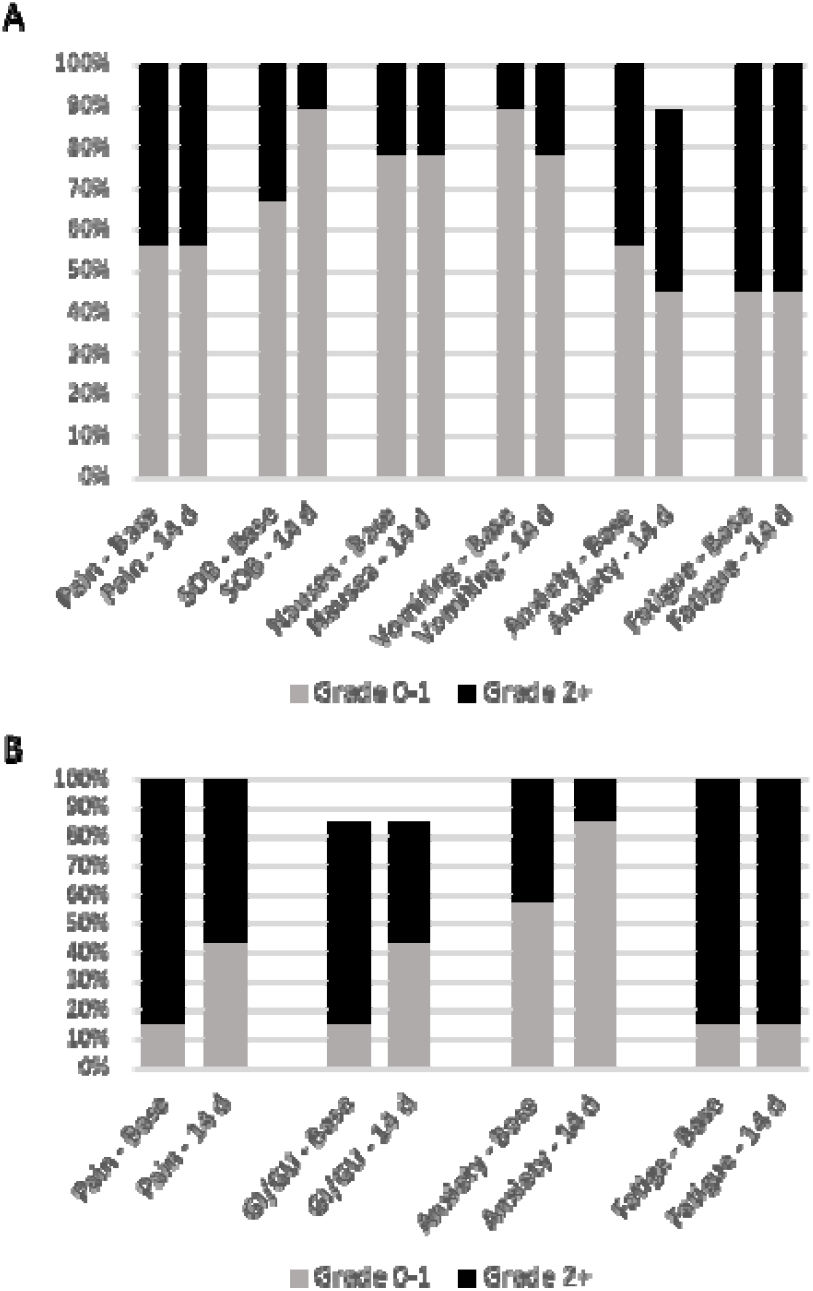
<u>Proportion of patients reporting PRO-CTCAE grade 0-1 and grade 2+ at baseline and 14 days.</u> A site-specific (thorax, gastrointestinal/pelvic) PRO-CTCAE inventory was assessed at each time point per protocol. 3A shows composite scores for pain, shortness of breath (SOB), nausea, vomiting, Anxiety, and fatigue for patients with thoracic tumors. 3B shows composite scores for pain, gastrointestinal (GI) and genitourinary symptoms (GU), anxiety, and fatigue.

Median PROMIS T-scores are shown in Table 3. These demonstrate that, at baseline, patients had more anxiety (median=55.7), depression (median=55.4), pain interference (median=61.3), and less physical global health (median=32.4) and physical function (median=34.5) than healthy individuals. At 14 days after treatment, patients improved on all measures. While data from only seven patients were available at 90 days, median anxiety (40.5), physical global health (39.2), depression (41.0), pain interference (41.6), and physical function (43.3) were all improved.

**Table 3:** Median (range), of PROMIS-QOL inventory T scores were assessed at baseline and each follow up time point, as available.

|  | ***Baseline (N=19)*** | ***14-day (N=15)*** | ***30-day (N=11)*** | ***90-day (N=7)*** |
| --- | --- | --- | --- | --- |
| ***Anxiety*** | 55.7 (40.5–72.5)* | 40.5 (40.5–67.6) | 48.4 (40.5–62.4) | 40.5 (40.5–52.2) |
| ***Physical global health*** | 32.4 (25.9–57.8) | 39.2 (25.9–57.8) | 39.2 (25.9–57.8) | 39.2 (32.4–57.8) |
| ***Depression*** | 55.4 (41–73.3) | 41 (41–71.6) | 41 (41–60.5), 11 | 41 (41–55.3)‡ |
| ***Pain interference*** | 61.3 (41.6–75.6) | 57.5 (41.6–75.6) | 54.8 (41.6–66.7)+ | 41.6 (41.6–59.7)§ |
| ***Physical function*** | 34.5 (23.5–61.5) | 37.5 (23.5–61.5) | 39.8 (27–61.5) | 43.3 (30.1–61.5) |

\* One patient did not complete the inventory (N=18)
+ One patient did not complete the inventory (N=17)
‡ Two patients did not complete the inventory (N=5)
§ One patient did not complete the inventory (N=6)

### Imaging Response

Thirteen patients with 14 tumors had diagnostic imaging at least 30 days following Lattice SBRT that were included for analysis. The tumor volume at each imaging time point, normalized by the original GTV volume, for all evaluable tumors is depicted in Supplementary Figure 2. The first follow up scan was a median of 1.8 months following Lattice SBRT, and all evaluable GTVs had decreased in size (median 24.4% decrease, range: 2.4–91.7%). Eleven patients (12 tumors) had a second follow up scan at a median of 4.5 months, and all but one tumor continued to shrink, with one tumor growing slightly (median 47.4% decrease, range: −5.3– 96.1%).

### Plasma Cytokine Response

Overall, plasma cytokine concentrations trended downward immediately following irradiation and at 14-day follow-up, but no changes among collection time points reached statistical significance (Supplementary Figure 3, adjusted *p*-values 0.237 - 1 by pairwise Wilcoxon rank sum test). Some plasma cytokine concentrations correlated across cytokine classes in pre-treatment samples, such as Th1-secreted cytokines IL-2, IL-10, INF-gamma, and TNF-alpha (Supplementary Figure 4A). Weaker associations among cytokines were generally lost immediately following Lattice irradiation (Supplementary Figure 4B), with the notable exception of eotaxin. The entire plasma cytokine profile is perturbed at 14-day follow-up (Supplementary Figure 4C), but pre-treatment associations start to be restored at 30-day follow-up (Supplementary Figure 4D).

## Discussion

This prospective phase I study found Lattice SBRT is safe to deliver as palliative radiotherapy for patients with very large tumors, including those who receive systemic therapy before or after radiation. We did not observe grade 3+ toxicities probably or definitely attributable to Lattice SBRT, and PROs suggest that the acute effects of Lattice SBRT did not add significant toxicity or harm QoL. There was only one instance of grade 2 radiation pneumonitis in patients with thoracic tumors (N=10). This patient had a large solitary fibrous tumor of the right upper lobe of the lung, abutting the mediastinum. While his Lattice SBRT plan met protocol constraints, it is of interest to note that the patient received immune checkpoint inhibitor therapy alone approximately 8 months prior to commencement of radiation. It is possible that he had higher propensity to experience radiation pneumonitis than other patients.^22^

Few studies have evaluated SBRT for tumors larger than 5 cm and all are retrospective.^6–8^ However, on average, the patients treated in these series had much smaller tumors than those treated in the current study with a reported median largest dimension 5.6 cm (versus 10.5 cm) or median GTV of 52.9 cc (versus 783 cc) with a reported max 597.8 cc (versus 3713 cc). Despite this difference, this study was associated with similar or lower toxicity rates compared with the prior work. For example, the largest study completed by Verma and colleagues reported grade 3+ toxicities in 5% of patients with nine cases (10%) of grade 2+ radiation pneumonitis.^8^ Two other retrospective series show a similar grade 3+ toxicity rates of 10%^6^ and 13%^7^ with grade 2+ pneumonitis rates of 12%^6^ and 7%^7^. It appears that our spatially fractionated SBRT approach offers a similar or better acute toxicity profile to conventional SBRT even for tumors significantly larger than those reported in the literature.

The exploratory analyses suggest that Lattice SBRT is efficacious as a palliative treatment for patients with large tumors and produces a measurable change in cytokine/chemokine levels. PRO-CTCAE data show that patients did not experience increased toxicity with Lattice SBRT except one patient each who experienced worse anxiety and nausea. PROMIS inventories demonstrated that patients had improved physical function, pain, global health, anxiety, and depression at 14 days after treatment and this improvement was preserved at 90 days. Analysis of GTVs demonstrated clinically significant shrinkage (median 24.4% by volume) within a median of 81 days following Lattice SBRT despite the fact that patients had very large tumors from cancers that are traditionally considered radioresistant (i.e. sarcomas, NSCLC). Further, 36% of evaluable tumors (N=11) had greater than 80% reduction of volume by the second imaging time point 3-6 months after treatment. The changes in cytokine/chemokine levels pre-/post-Lattice SBRT indicates that this approach might induce a measurable immune-mediated response, similar to conventional SBRT. It is difficult, however, to extrapolate from our findings to identify a specific leukocyte subset given the number of patients analyzed, heterogeneity of this population, and pre-/post-Lattice SBRT systemic therapies. Nevertheless, we believe that this preliminary data supports further investigation in future clinical trials of Lattice SBRT.

This study has important limitations. First, toxicity was only prospectively evaluated for 90 days. As SBRT can be associated with significant late toxicity, it is vital to evaluate for late toxicity associated with Lattice SBRT. Second, patients in this study reported substantial baseline toxicity, and PROMIS measures demonstrate significantly worse QoL compared to healthy individuals. Only half of patients were able to complete trial follow up and 4 died within 90 days of completing Lattice SBRT. Third, the study evaluated a highly heterogeneous cohort in order to evaluate the safety of a promising novel radiotherapy option for patients that are otherwise treated with a range of radiotherapy doses and fractionations, other local therapy options, or supportive care. The ongoing prospective phase II study of Lattice SBRT that is enrolling patients in to pre-specified thoracic, abdomen, pelvis, and sarcoma sub-groups will address some of these limitations (NCT 04553471).

## Data Availability

The data are stored locally, please contact the corresponding author to discuss.

## Supplementary Materials

### Methods for cytokine/chemokine analysis

Whole blood was collected in EDTA-treated tubes and processed to plasma, which was subsequently flash-frozen and stored at −80°C. Frozen plasma was slowly thawed to room temperature, and co-incubated with samples from the Invitrogen Cytokine & Chemokine 34-Plex

Human ProcartaPlex™ Panel 1A, as well as immunoconjugates against interferon beta, and programmed death-ligand 1 (Thermo Fisher Scientific, Waltham). Antibody/analyte complexes were run on a Luminex™ FLEXMAP 3D (Luminex, Austin). Quantitative Luminex data were analyzed with BelysaTM software (Millipore-Sigma, St. Louis).

**Supplementary Figure 1.**
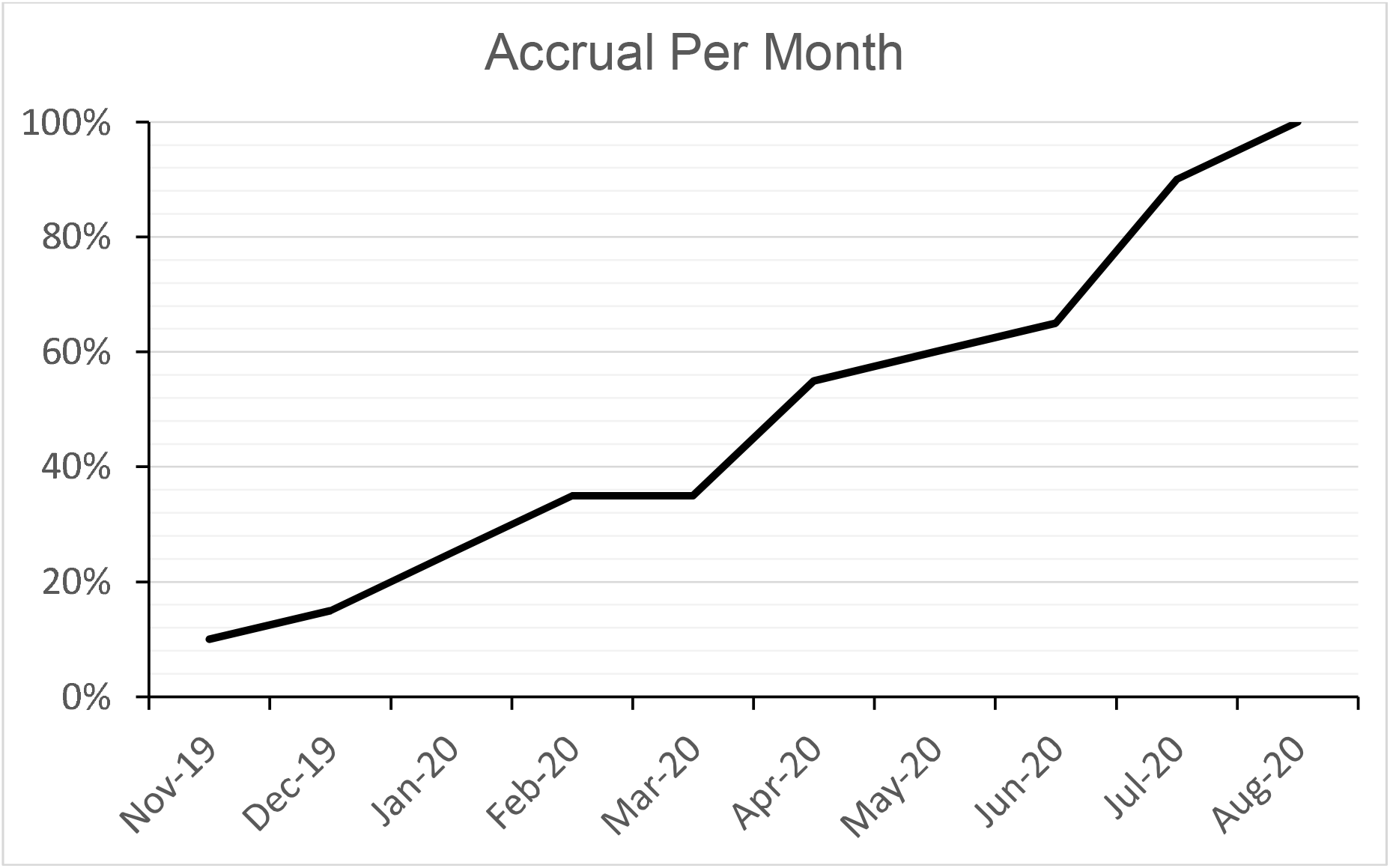

**Supplementary Figure 2:**
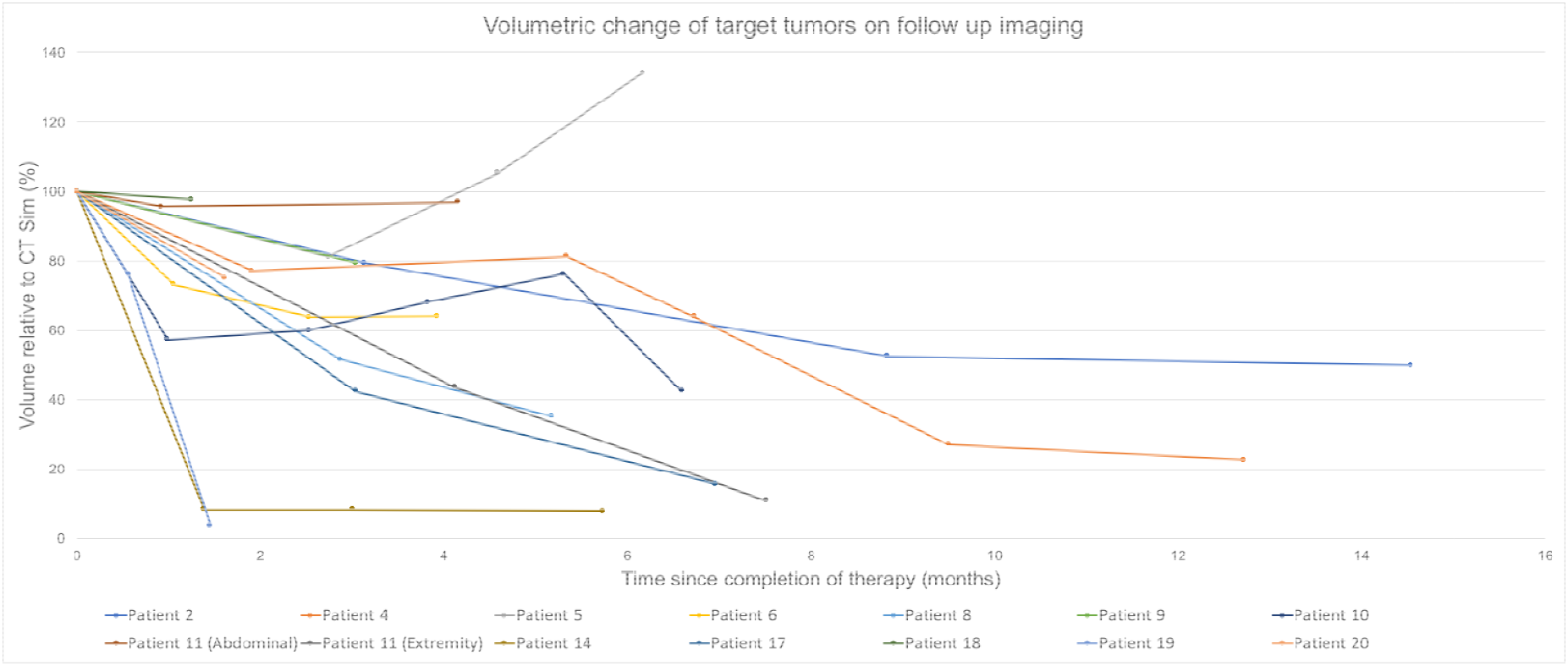
Change in GTV on follow up imaging. Diagnostic images were fused with CT Sim data in the TPS to contour tumors and measure volumetric change after completion of Lattice SBRT. Change relative to baseline GTV_2000 is shown against time (in months).

**Supplementary Figure 3:**
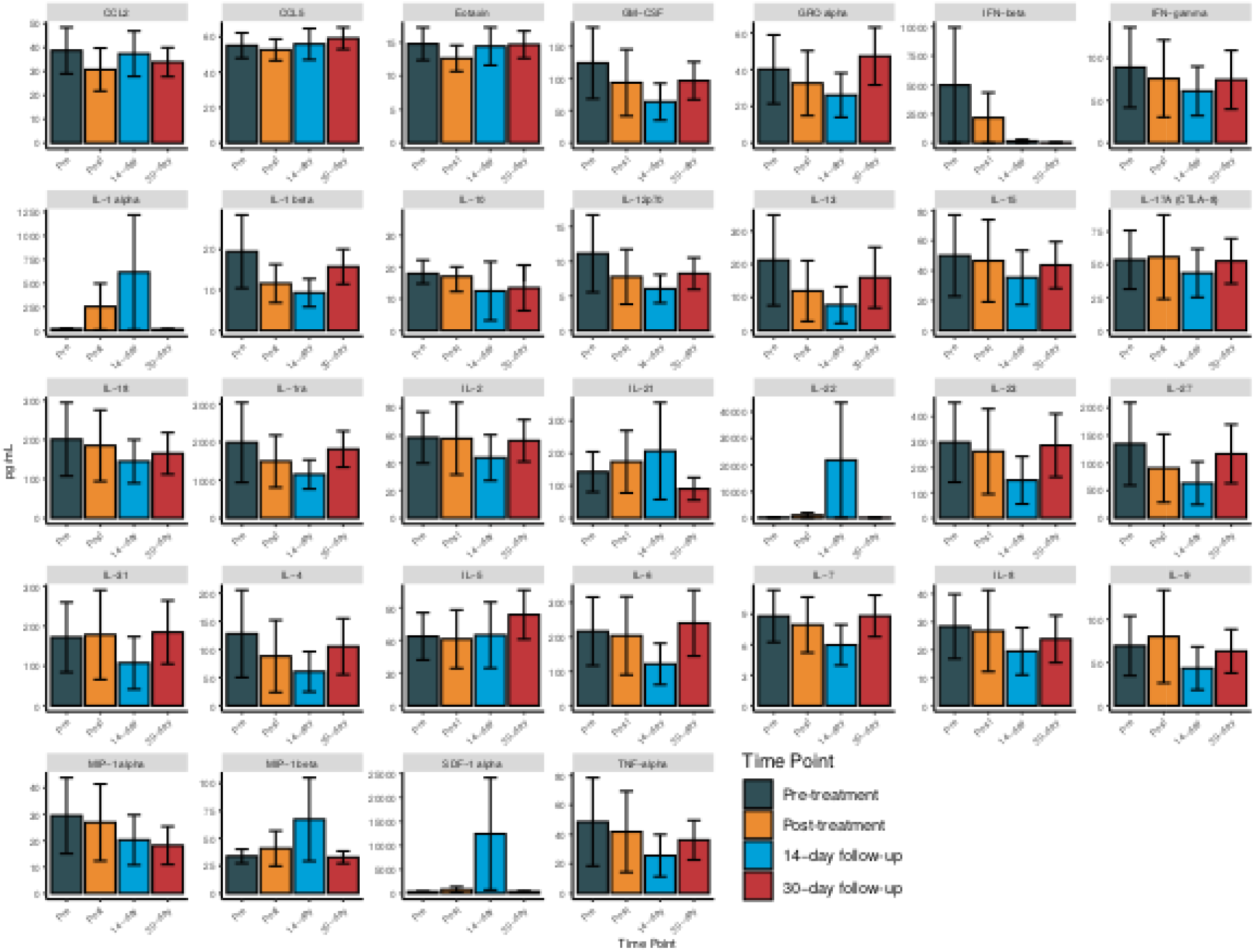
Mean plasma cytokine concentration at each time point. Bars are standard error of measurements for all samples available at each time point.

**Supplementary Figure 4:**
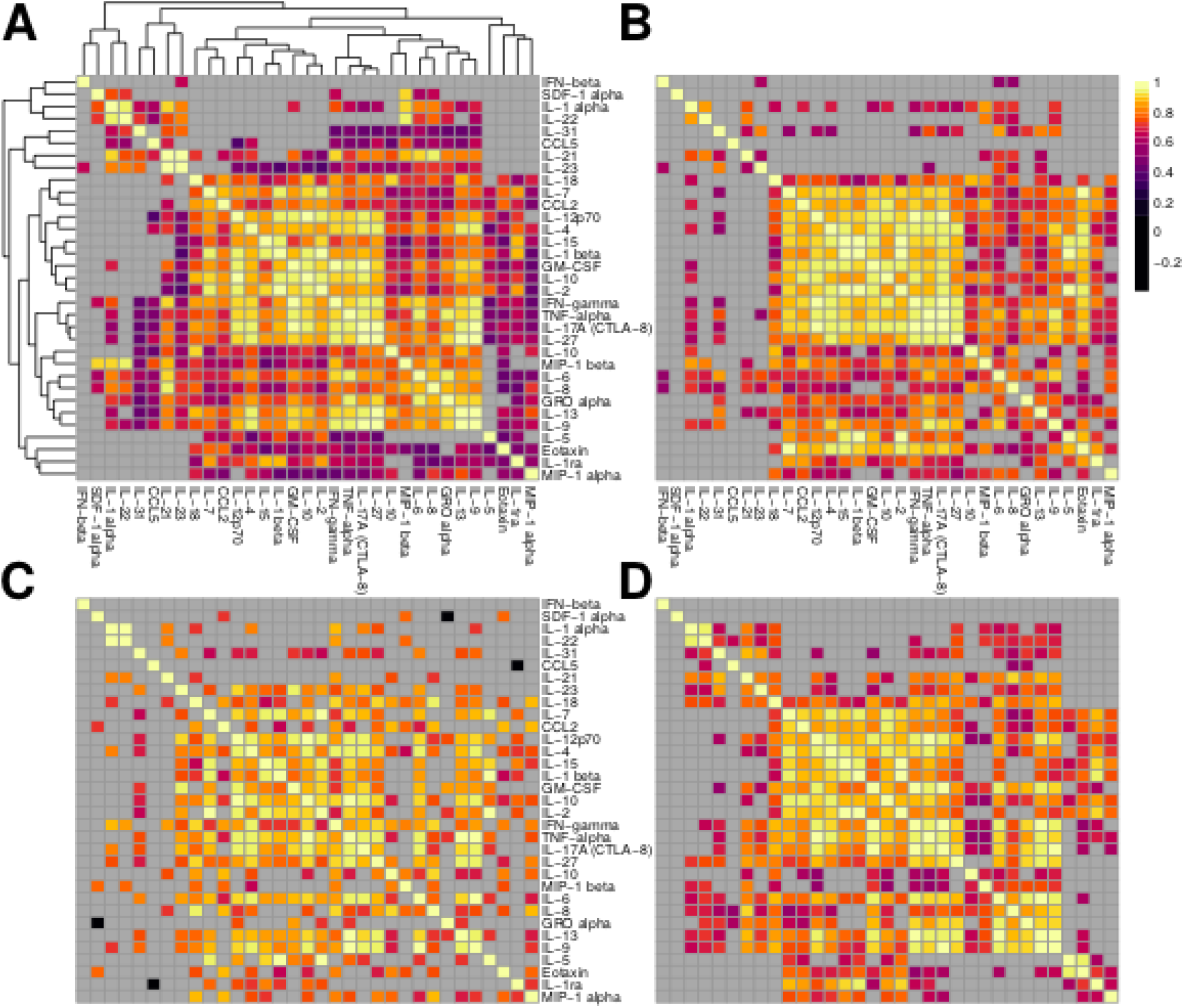
Spearman correlation of plasma cytokines at each time point. Heatmaps of spearman rank-order correlations among cytokines at A) pre-treatment, B) post-treatment, C) 14-day follow-up, and D) 30-day follow-up. Only correlations with *p*-value < 0.05 are shown. Panel A clustering is arranged by Ward’s method. Panels B-D retain the clustering order of panel A for comparison.

**Supplementary Table 1.**
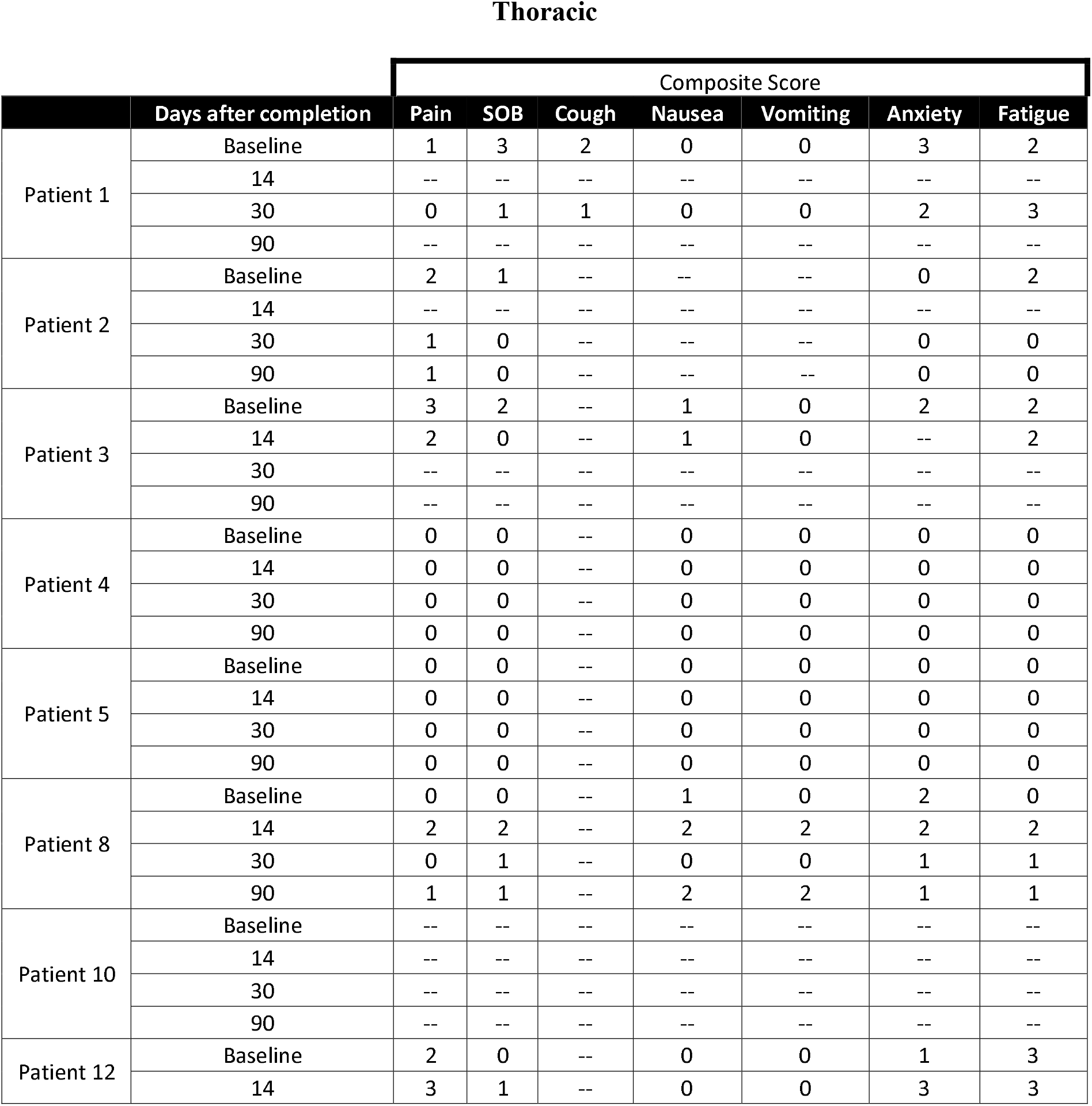
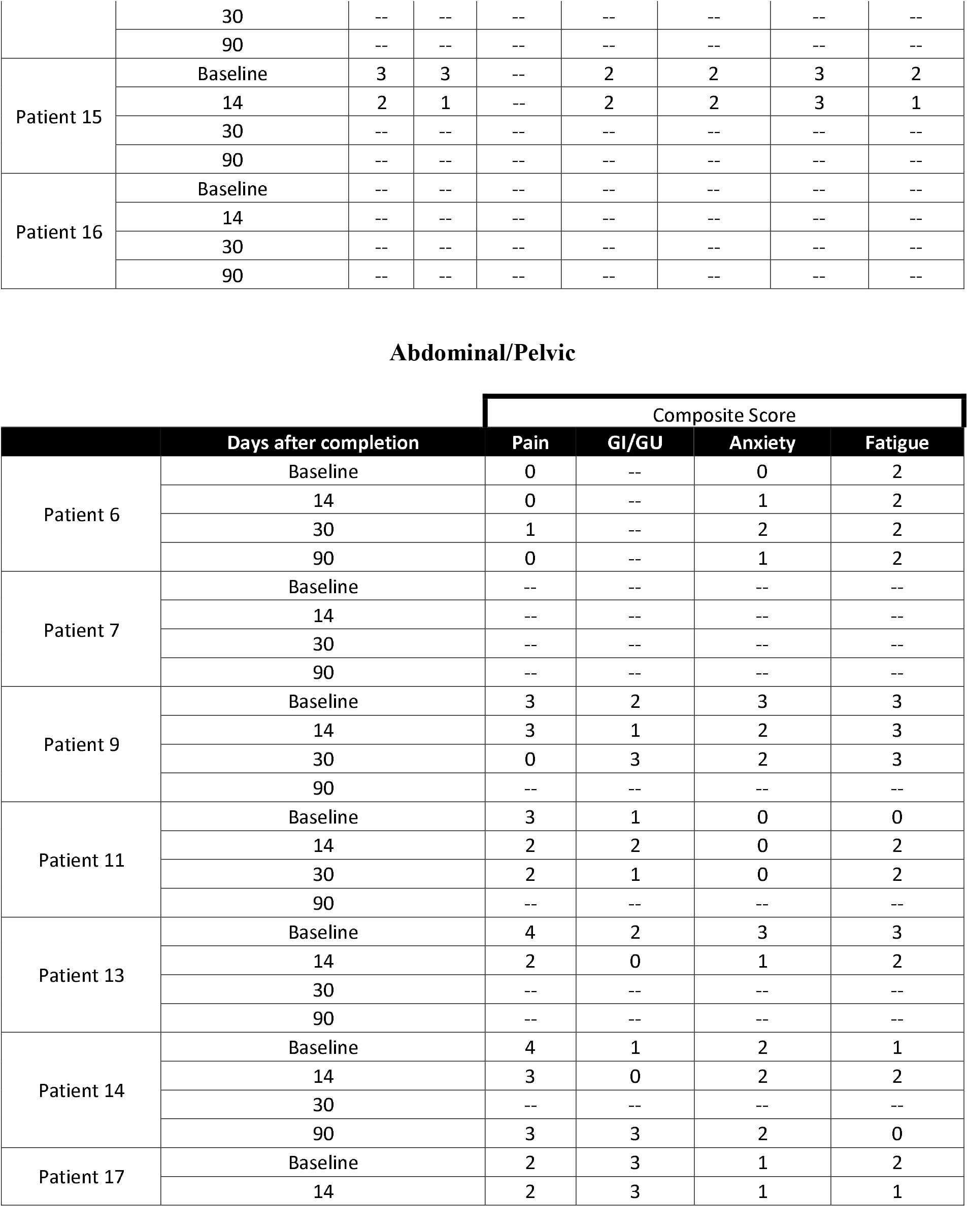
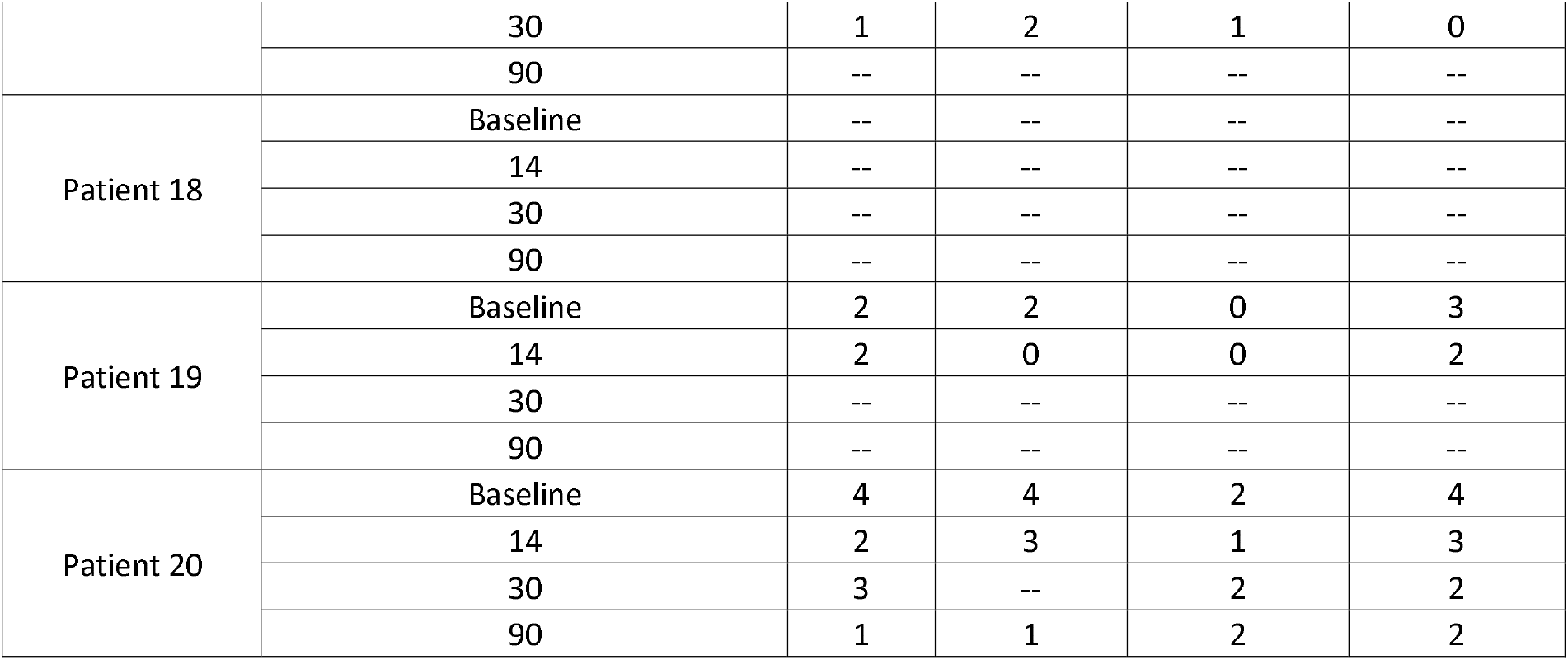
PRO-CTCAE data for all patients at all follow-up time points. Patients are grouped by treatment area (thoracic vs abdominal/pelvic). An entry of -- indicates that the PRO was not collected at that time point due to death, loss to follow-up, or withdrawal from study.

